# Assessment of ability of AlphaMissense to identify variants affecting susceptibility to common disease

**DOI:** 10.1101/2023.11.22.23298908

**Authors:** David Curtis

## Abstract

An important issue pertinent to the analysis of sequence data to detect association between rare variants in a gene and a given phenotype is the ability to annotate nonsynonymous variants in terms of their likely importance as affecting protein function. While a number of software tools attempt to do this, AlphaMissense was recently released and was shown to have good performance using benchmarks based on variants causing severe disease and on functional assays. Here, we assess the performance of AlphaMissense across 18 genes which had previously demonstrated association between rare coding variants and hyperlipidaemia, hypertension or type 2 diabetes. Ability to detect association, expressed as the signed log p value (SLP) was compared between AlphaMissense and 43 other annotation methods. The results demonstrated marked variability between genes regarding the extent to which nonsynonymous variants contributed to evidence for association and also between the performance of different methods of annotating the nonsynonymous variants. Although AlphaMissense produced the highest SLP on average across genes, it produced the maximum SLP for only 4 genes. For some genes, other methods produced a considerably higher SLP and there were examples of genes where AlphaMissense produced no evidence for association while another method performed well. The marked inconsistency across genes means that it is difficult to decide on an optimal method of analysis of sequence data. The fact that different methods perform well for different genes suggests that if one wished to use sequence data for individual risk prediction then gene-specific annotation methods should be used. It would be desirable to have the ability to recognise characteristics of a gene which could facilitate the selection of an annotation method which would best discriminate variants of interest within that gene.

This research has been conducted using the UK Biobank Resource.

## Introduction

As large exome-sequenced datasets become available it has become possible to detect gene-level associations between the burden of extremely rare coding variants and a variety of phenotypes (Backman et al., 2021; Wang et al., 2021). Typically, tests for association involve considering together variants falling into a particular category based on their predicted effect. Variants expected to completely disrupt function of a gene, consisting of stop gained, frameshift and splice site variants are jointly termed loss of function (LOF) or protein truncating variants (PTV) and when considered jointly it is usually the case that this category of variant is associated with the largest effect size. While nonsynonymous variants may also have effects, the nature and magnitude of these effects is likely to be heterogeneous and if all nonsynonymous variants are considered to form a single category then the average estimated effect will naturally be smaller than that of the variants having the largest effect sizes. Rare variant association studies which simply consider all nonsynonymous variants jointly have yielded informative results (Sazonovs et al., 2022). However a more widely used approach is to use some form of secondary annotation method which attempts to distinguish those nonsynonymous variants which are more likely to have a biological effect and applying such approaches may allow one to demonstrate that those nonsynonymous variants predicted to be most impactful are indeed the ones which show association with a phenotype (Singh and The Schizophrenia Exome Meta-Analysis (SCHEMA) Consortium, 2022).

A large number of methods are available to carry out such secondary annotations and we have recently assessed their relative performance (Curtis, 2022). Since then a new method, AlphaMissense, has been released with the aim of recognising whether a nonsynonymous variant observed in a patient is or is not likely to be pathogenic (Cheng et al., 2023). The report of this study also discusses at length the various issues involved in attempting to interpret the likely effects of nonsynonymous variants. The AlphaMissense prediction is based on machine learning approaches to assimilate information about the protein structural context and about evolutionary conservation to generate a score reflecting likely pathogenicity. It was demonstrated to perform well on benchmarks derived from clinically identified variants as well as from multiplexed assays of variant effect (MAVEs).

Although one might hope that the same classification methods used to identify single variants causing severe disease might also be helpful in attempting to discriminate those variants increasing risk of common phenotypes, this is not necessarily the case. For example, when using PolyPhen-2 it is recommended that the version trained on HumVar be used to assist the diagnosis of Mendelian disorders while the version trained on HumDiv should be used to evaluate rare alleles for complex genotypes (Adzhubei et al., 2010). This consideration means that it would be helpful to assess the extent to which AlphaMissense could assist as an annotation tool in the context of large case control studies of exome sequenced datasets aiming to identify genes influencing the risk of common phenotypes.

The aim of the present study is to compare the performance of AlphaMissense with other annotation methods in terms of their ability to produce evidence for association between a gene and a common, clinically relevant phenotype. Such associations were previously established using weighted burden analyses, in which different variants within a gene were weighted differentially according to their annotation and rarity. In the present analyses, variants are weighted for rarity and then the contributions to evidence for association are examined separately for AlphaMissense and a number of other annotation methods.

## Methods

The methods used closely followed those described in the previous study exploring different annotation and weighting schemes, and the description is partly repeated here for the convenience of the reader (Curtis, 2022).

The UK Biobank Research Analysis Platform was used to access the Final Release Population level exome OQFE variants in PLINK format for 469,818 exomes which had been produced at the Regeneron Genetics Center using the protocols described here: https://dnanexus.gitbook.io/uk-biobank-rap/science-corner/whole-exome-sequencing-oqfe-protocol/protocol-for-processing-ukb-whole-exome-sequencing-data-sets (Backman et al., 2021). UK Biobank had obtained ethics approval from the North West Multi-centre Research Ethics Committee which covers the UK (approval number: 11/NW/0382) and had obtained written informed consent from all participants. The UK Biobank approved an application for use of the data (ID 51119) and ethics approval for the analyses was obtained from the UCL Research Ethics Committee (11527/001). To obtain 20 population principal components reflecting ancestry, version 2.0 of plink (https://www.cog-genomics.org/plink/2.0/) was run with the options --*maf 0*.*1 --pca 20 approx* (Chang et al., 2015; Galinsky et al., 2016).

To assess overall evidence for gene-wise associations with different phenotypes, weighted burden analyses had previously been carried out using the SCOREASSOC and GENEVARASSOC programs (Curtis, 2016). Attention was restricted to rare variants with minor allele frequency (MAF) <= 0.01 in both cases and controls. As previously described, variants were weighted by overall MAF so that variants with MAF = 0.01 were given a weight of 1 while very rare variants with MAF close to zero were given a weight of 10, with a parabolic function used to assign weights with intermediate MAFs (Curtis, 2020). Additionally each variant was annotated with the Variant Effect Predictor (VEP), SIFT and PolyPhen SIFT (Adzhubei et al., 2013; Kumar et al., 2009; McLaren et al., 2016). A weight was assigned according to this annotation and the overall weight for each variant consisted of the frequency weight multiplied by the annotation weight. For each subject and each gene, the weights for the variants carried by the subject were summed to provide an overall weighted burden score. Regression modelling was done to calculate the likelihood for the phenotype data given covariates consisting of sex and the first 20 principal components and then the likelihood was recalculated for the model additionally incorporating the weighted burden score. Twice the natural log of the ratio of these likelihoods is a likelihood ratio statistic taken to be distributed as a chi-squared statistic with 1 degree of freedom. The evidence for association is summarised as the signed log p value (SLP) taken as the log base 10 of the p value and given a positive sign if there is a positive correlation between the weighted burden score and the phenotype.

For the present study variant annotation was performed in two stages. First, a primary categorisation was made using VEP, which uses information based on the reference sequence and coordinates of known transcripts to report findings such as whether variants occur within exons, if so whether they change amino acid sequence, etc (McLaren et al., 2016). For purposes of the present analyses, variants predicted to have a similar kind of effect were grouped together so that, for example, stop gained, frameshift and essential splice site variants were all treated as LOF. The full list of annotations as reported by VEP and the category they were assigned to is shown in Table 1, along with the weights which were used for the previous weighted burden analyses, which had been arbitrarily assigned based on expectations of the likely biological importance of each annotation. Each of the annotation categories was then used to generate a separate burden score, so that for example the burden score relating to the category LOF for a subject would consist of the number of LOF variants carried by that subject, each multiplied by the weight according to allele frequency as described above.

**Table 1.** Table showing annotations produced by VEP, the weights assigned to them for the previous weighted burden analyses and the categories they were assigned to for the current analyses. Annotations marked as unused were not applied to any of the variants in the genes studied.

| **VEP annotation** | **Weight** | **Category** |
| --- | --- | --- |
| intergenic_variant | 0 | Unused |
| feature_truncation | 0 | IntronicEtc |
| regulatory_region_variant | 0 | IntronicEtc |
| feature_elongation | 0 | IntronicEtc |
| regulatory_region_amplification | 1 | IntronicEtc |
| regulatory_region_ablation | 1 | IntronicEtc |
| TF_binding_site_variant | 1 | IntronicEtc |
| TFBS_amplification | 1 | IntronicEtc |
| TFBS_ablation | 1 | IntronicEtc |
| downstream_gene_variant | 0 | IntronicEtc |
| upstream_gene_variant | 0 | IntronicEtc |
| non_coding_transcript_variant | 0 | IntronicEtc |
| NMD_transcript_variant | 0 | IntronicEtc |
| intron_variant | 0 | IntronicEtc |
| non_coding_transcript_exon_variant | 0 | IntronicEtc |
| 3_prime_UTR_variant | 1 | ThreePrime |
| 5_prime_UTR_variant | 1 | FivePrime |
| mature_miRNA_variant | 5 | Unused |
| coding_sequence_variant | 0 | Unused |
| synonymous_variant | 0 | Synonymous |
| stop_retained_variant | 5 | Synonymous |
| incomplete_terminal_codon_variant | 5 | Unused |
| splice_region_variant | 1 | SpliceRegion |
| protein_altering_variant | 5 | ProteinAltering |
| missense_variant | 5 | ProteinAltering |
| inframe_deletion | 10 | InDelEtc |
| inframe_insertion | 10 | InDelEtc |
| transcript_amplification | 10 | InDelEtc |
| start_lost | 10 | ProteinAltering |
| stop_lost | 10 | ProteinAltering |
| frameshift_variant | 100 | LOF |
| stop_gained | 100 | LOF |
| splice_donor_variant | 100 | LOF |
| splice_acceptor_variant | 100 | LOF |
| transcript_ablation | 100 | LOF |

In order to obtain secondary annotations using AlphaMissense for all nonsynonymous variants, VEP was run with the options *b --canonical –regulatory --plugin AlphaMissense* (Cheng et al., 2023).This produces two AlphaMissense annotations, a raw score and a categorisation of likely pathogenic, likely benign or ambiguous. These three categories were converted to numerical scores of 2, 0 or 1 respectively. To obtain secondary annotations for other predictors, dbNSFP v4 was used (Liu et al., 2020). For the nonsynonymous and splice site variants listed in dbNSFP v4, secondary annotation scores were obtained consisting of the rank scores for a variety of different prediction and conservation methods. For each secondary annotation for a variant, the annotation score was then multiplied by the weight based on allele frequency. Thus, a subject’s overall score for the SIFT annotation would consist of the sum of all the SIFT rank scores of the variants carried by that subject, with the score for each variant also each being weighted according to allele frequency. For ease of processing, special characters in dbNSFP annotation names were replaced, for example GERP++ was changed to GERPPP. A total of 43 such scores were used, as presented below as and as detailed at http://database.liulab.science/dbNSFP.

The genes selected for this study consisted of those which had previously produced exome-wide significant results in weighted burden analyses using phenotypes of hypertension, hyperlipidaemia and type 2 diabetes (Curtis, 2023a, 2023b, 2023c). These genes and phenotypes are listed in Table 2. For each phenotype, a mixture of self-report, recorded diagnoses and medication reports was used to designate a set of participants as cases, with all other participants taken to be controls. There were a total of 469,818 exome-sequenced UK Biobank participants, of whom 167,127 were designated cases for hypertension, 106,091 for hyperlipidaemia and 33,629 for type 2 diabetes. As noted in the legend for Table 2, for some of these genes the original SLPs obtained were negative, indicating that variants impairing the function of these genes were protective and were associated with lower risk of developing the clinical phenotype. For the purpose of the current study, in order to make it easier to interpret the results for these genes alongside the others, the phenotype of interest for these genes is taken to be “being a control”, meaning that all variants associated with the phenotype would tend to generate positive SLPs.

**Table 2.** List of genes used for these analyses along with the SLP obtained in the original analyses with the corresponding phenotype (Curtis, 2023a, 2023b, 2023c). Variants which impaired functioning of *NPC1L1, PCSK9, ANGPTL3* and *APOC3* were found to be protective against hyperlipidaemia so for convenience the phenotype of interest is stated to be “Not hyperlipidaemia”. Likewise, variants impairing functioning of *INPPL1* and *DBH* are protective against hypertension.

| Phenotype | Gene symbol | Gene name | SLP |
| --- | --- | --- | --- |
| Hyperlipidaemia | *LDLR* | Low Density Lipoprotein Receptor | 156.81 |
| Hyperlipidaemia | *ABCG5* | ATP Binding Cassette Subfamily G Member 5 | 6.95 |
| Not hyperlipidaemia | *NPC1L1* | NPC1 Like Intracellular Cholesterol Transporter 1 | 7.60 |
| Not hyperlipidaemia | *PCSK9* | Proprotein Convertase Subtilisin/Kexin Type 9 | 43.57 |
| Not hyperlipidaemia | *ANGPTL3* | Angiopoietin Like 3 | 12.68 |
| Not hyperlipidaemia | *APOC3* | Apolipoprotein C3 | 13.19 |
| Hypertension | *DNMT3A* | DNA Methyltransferase 3 Alpha | 14.20 |
| Hypertension | *FES* | FES Proto-Oncogene, Tyrosine Kinase | 9.92 |
| Hypertension | *SMAD6* | SMAD Family Member 6 | 6.02 |
| Hypertension | *NPR1* | Natriuretic Peptide Receptor 1 | 7.98 |
| Hypertension | *GUCY1A1* | Guanylate Cyclase 1 Soluble Subunit Alpha 1 | 9.13 |
| Hypertension | *ASXL1* | ASXL Transcriptional Regulator 1 | 8.35 |
| Not hypertension | *INPPL1* | Inositol Polyphosphate Phosphatase Like 1 | 7.09 |
| Not hypertension | *DBH* | Dopamine Beta-Hydroxylase | 9.71 |
| Type 2 diabetes | *GCK* | Glucokinase | 32.11 |
| Type 2 diabetes | *HNF4A* | Hepatocyte Nuclear Factor 4 Alpha | 9.39 |
| Type 2 diabetes | *HNF4A* | Hepatocyte Nuclear Factor 4 Alpha | 7.98 |
| Type 2 diabetes | *GIGYF1* | GRB10 Interacting GYF Protein 1 | 7.58 |

To gain an understanding of the relationships between the different annotation methods, a correlation matrix was produced of all the secondary annotation scores across all the nonsynonymous variants in all these genes and this matrix was visualised using the correl package in R (Makowski et al., 2020; R Core Team, 2014).

In order to assess the contributions of each different category of variant to the evidence for association, a logistic regression analysis was performed separately on the weighted burden score for each of the primary categories, with population principal components and sex being included as covariates. The Wald statistic was then used to obtain an SLP for each variant category for each gene and these were tabulated and compared.

Similar analyses were performed for secondary annotations obtained from AlphaMissense and dbNSFP, except that for these analyses the weighted burden score produced by the ProteinAltering category was included as an additional covariate. This is because the overall burden for each of these secondary annotations would depend on the total the number of nonsynonymous variants each subject carries and the purpose of these analyses is to assess the relevant performance of the different secondary annotation methods to distinguish the effect of different nonsynonymous variants. Again, the Wald statistic was used to obtain SLPs for each secondary annotation and these were tabulated and compared.

Data manipulation and statistical analyses were performed using GENEVARASSOC, SCOREASSOC and R (Curtis, 2020, 2016; R Core Team, 2014).

## Results

Figure 1 shows a heatmap which illustrates the relative magnitude of the SLP produced by each variant category for each gene and the actual SLPs are shown in Table 3. From this it can be seen that for most genes the only variant categories to generate SLPs of a large magnitude were LOF and ProteinAltering. However for *ABCG5* and *ANGPTL3* the SpliceRegion category had large SLPs whereas for *LDLR* and *HNF4A* the InDelEtc category had large SLPs. For some genes both the LOF and ProteinAltering categories had large SLPs but for others only one category did. For example, for *HNF1A* the LOF category produced a much larger SLP than ProteinAltering did, whereas for *HNF41A* this situation was reversed and the ProteinAltering category produced a fairly large SLP whereas the LOF SLP was minimal. Thus it can be seen that there is no consistency across genes regarding which variant categories make the most substantial contribution to evidence for association, implying that no single scheme could be optimal for all genes.

**Figure 1.**
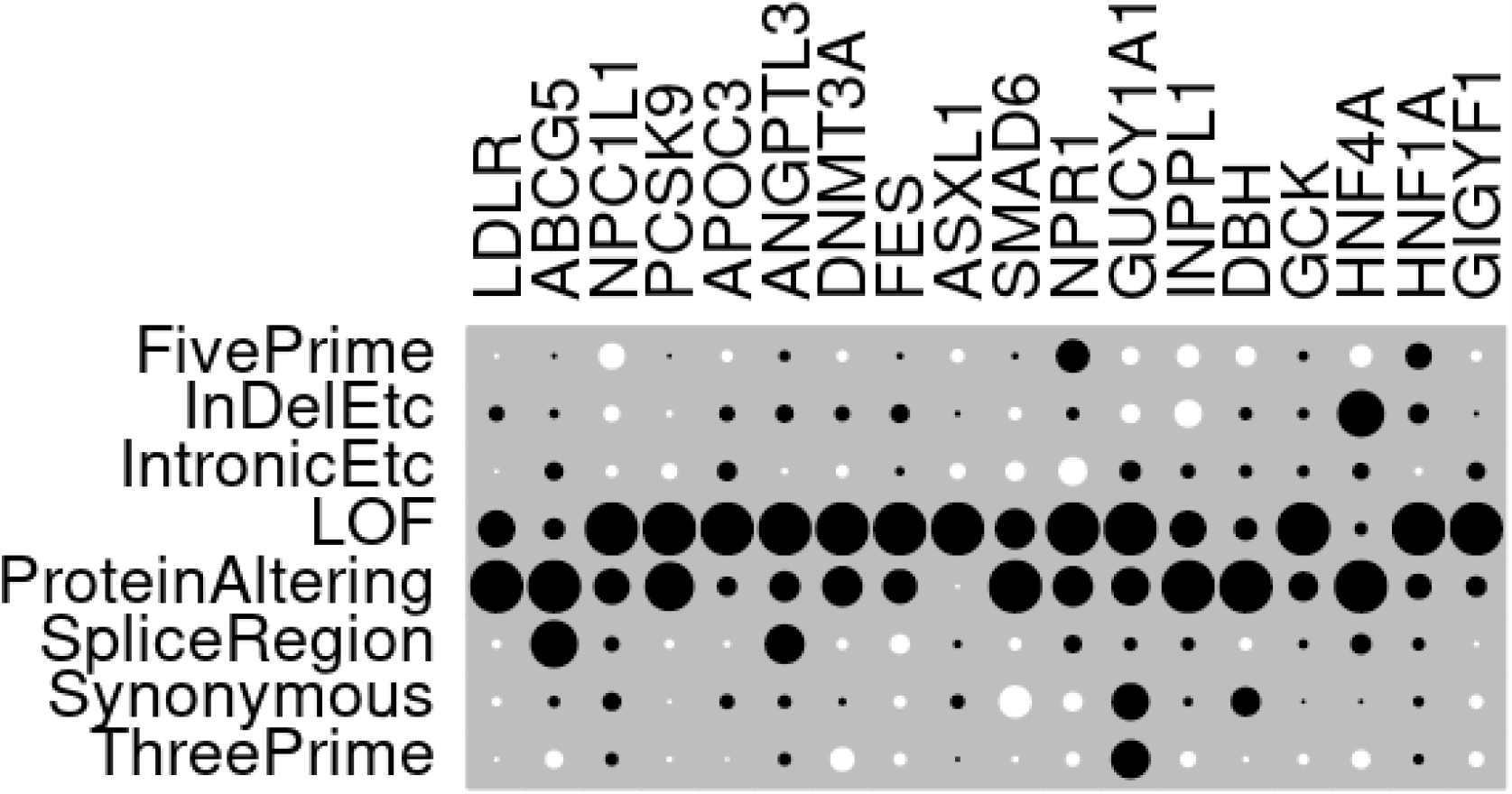
Heatmap of SLPs produced by each variant category for each gene. The sizes of the dots for each gene are proportional to the SLP for each variant category relative to the maximum category SLP obtained for that gene. White circles indicate negative SLPs.

**Table 3.** SLPs produced individually by each variant category for each gene, including sex and principal components as covariates. SLPs of 3 or more are shown in bold and SLPs of 6 or more in bold italics. The final column shows the mean SLP achieved by each category across all genes.

| Variant category | *LDLR* | *ABCG5* | *NPC1L1* | *PCSK9* | *APOC3* | *ANGPTL3* | *DNMT3A* | *FES* | *ASXL1* | *SMAD6* | *NPR1* | *GUCY1A1* | *INPPL1* | *DBH* | *GCK* | *HNF4A* | *HNF1A* | *GIGYF1* | Mean |
| --- | --- | --- | --- | --- | --- | --- | --- | --- | --- | --- | --- | --- | --- | --- | --- | --- | --- | --- | --- |
| FivePrime | -0.31 | 0.04 | -1.15 | 0.05 | -0.42 | 0.29 | -0.34 | 0.00 | -0.77 | 0.04 | 1.17 | -0.51 | -0.65 | -0.99 | 0.56 | -0.85 | 2.35 | -0.37 | -0.10 |
| InDelEtc | **4.82** | 0.18 | -0.41 | -0.17 | 0.81 | 0.83 | 0.68 | 0.50 | 0.09 | -0.20 | 0.15 | -0.62 | -0.99 | 0.38 | 0.75 | **4.12** | 1.38 | 0.05 | 0.69 |
| IntronicEtc | -0.03 | 0.82 | -0.21 | -1.36 | 1.31 | -0.08 | -0.44 | 0.09 | -1.09 | -0.49 | -0.83 | 0.75 | 0.25 | 0.39 | 0.89 | 0.47 | -0.15 | 1.11 | 0.08 |
| LOF | **30.87** | 1.15 | **5.15** | **17.57** | **10.88** | **8.36** | **9.70** | **4.49** | **14.11** | 2.29 | **3.09** | **5.36** | 1.80 | 1.36 | **21.42** | 0.27 | **10.42** | **10.83** | **8.84** |
| ProteinAltering | **65.60** | **7.80** | 2.28 | **14.78** | 1.34 | 2.68 | **5.47** | 1.84 | -0.04 | **4.09** | 1.75 | 2.67 | **3.98** | **7.82** | **6.23** | **5.42** | 2.25 | 1.50 | **7.64** |
| SpliceRegion | -1.35 | **6.07** | 0.36 | -0.39 | -0.12 | **4.57** | -0.32 | -0.55 | 0.23 | -0.18 | 0.32 | 0.28 | 0.21 | -0.36 | 0.50 | 0.73 | 0.49 | -0.06 | 0.58 |
| Synonymous | -1.45 | 0.29 | 0.56 | -0.06 | 0.71 | 0.44 | 0.14 | -0.20 | 0.84 | -1.45 | -0.37 | 2.58 | 0.09 | 2.25 | 0.03 | 0.02 | 0.33 | -0.62 | 0.23 |
| ThreePrime | -0.27 | -0.80 | 0.26 | -0.12 | -0.02 | 0.41 | -1.85 | -0.21 | 0.06 | -0.04 | -0.19 | 2.79 | -0.31 | -0.06 | -0.69 | -0.59 | 0.32 | -0.78 | -0.12 |

In order to gain insights into the relationships between the secondary annotations, pairwise correlation coefficients were obtained between all pairs across variants in all genes, comprising 10,567 nonsynonymous variants, and a heatmap illustrating these correlations is shown in Figure 2. The raw correlation coefficients themselves are tabulated in Supplementary Table 1. It can be seen that the AlphaMissense annotations are positively correlated with each other and with 15 other annotations, forming a block. There is then a second block comprising 8 annotations which are again positively correlated with each other but which show little correlation with any of the annotations forming the first block. Interestingly, the Mutation Predictor (MutPred) score is positively correlated with the annotations of the first block and somewhat negatively correlated with those in the second block (Li et al., 2009). Following these two blocks are a number of other annotations showing little in the way of correlation with any of the others. Notably, this list includes the CADD annotations, which are quite widely used but which somewhat surprisingly seem to pick up different variant characteristics than the other methods (Rentzsch et al., 2019).

**Figure 2.**
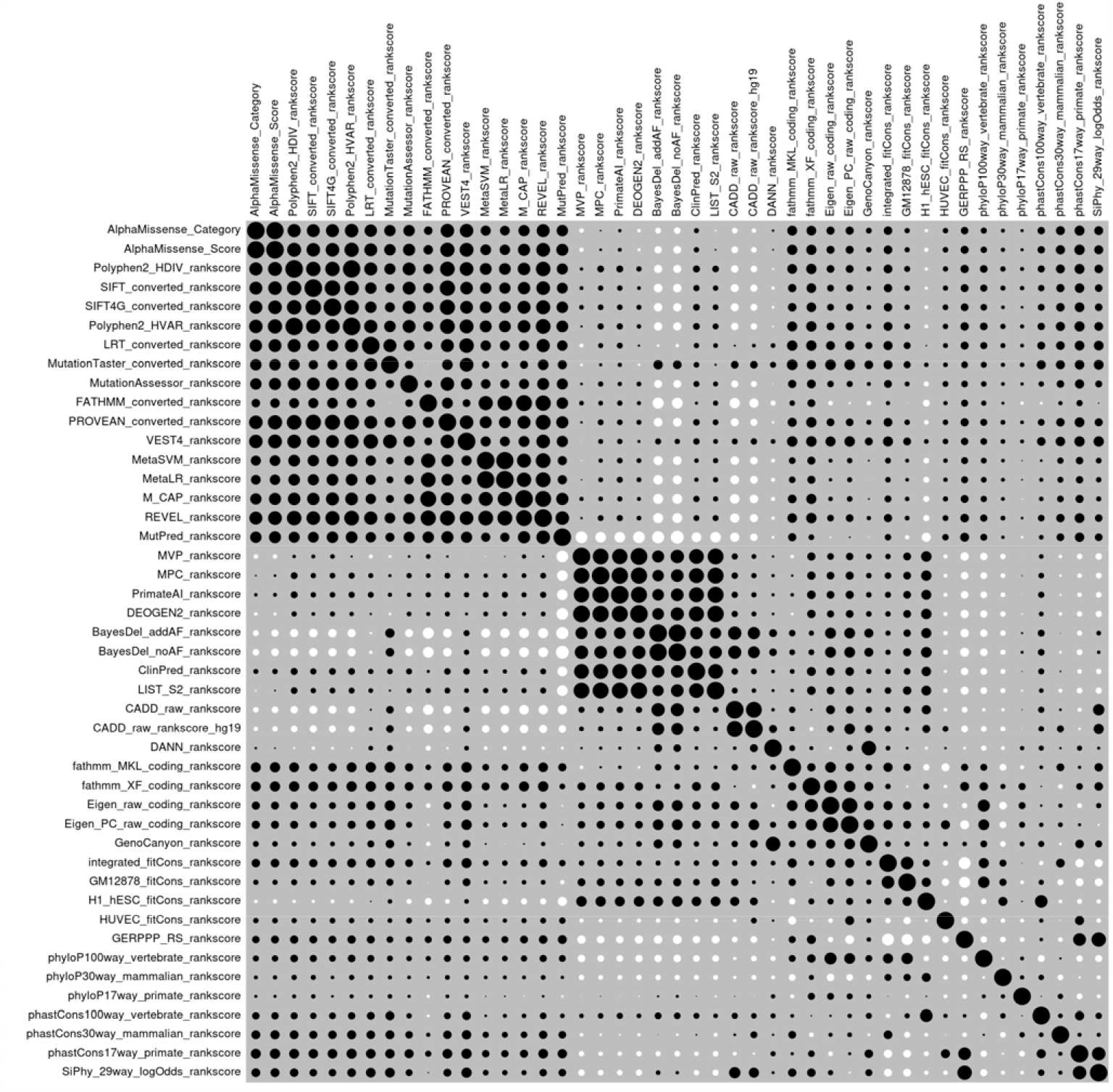
Plot of pairwise correlations between secondary annotations for the variants used in this study. Black circles indicate positive correlations and white circles negative correlations.

The relative performance of the different secondary annotations in terms of producing evidence for association is displayed as a heatmap in Figure 3 and the underlying SLPs are listed in Table 4. One thing of note is that there is considerable variability between genes as to the extent to which any of the secondary annotation methods produces evidence for association, as measured by the magnitude of the SLP. For some genes the methods are clearly quite effective. For example, *LDLR, PCSK9* and *GCK* all yield large SLPs for a variety of different annotations. Interestingly, although *APOC3* produced a negligible SLP of 1.34 for the ProteinAltering category taken as a whole, when these variants are annotated with MutationTaster they yield an SLP of 11.33 (Schwarz et al., 2014). Conversely, *ABCG5* produced SLP = 7.80 for the ProteinAltering category but none of the secondary annotation methods seems able to distinguish which variants within this category are more associated with risk and the maximum any of them produces is SLP = 2.25 for the SiPhy score, which is a conservation score based on comparison of human and mammalian genomes (Lindblad-Toh et al., 2011).

**Figure 3.**
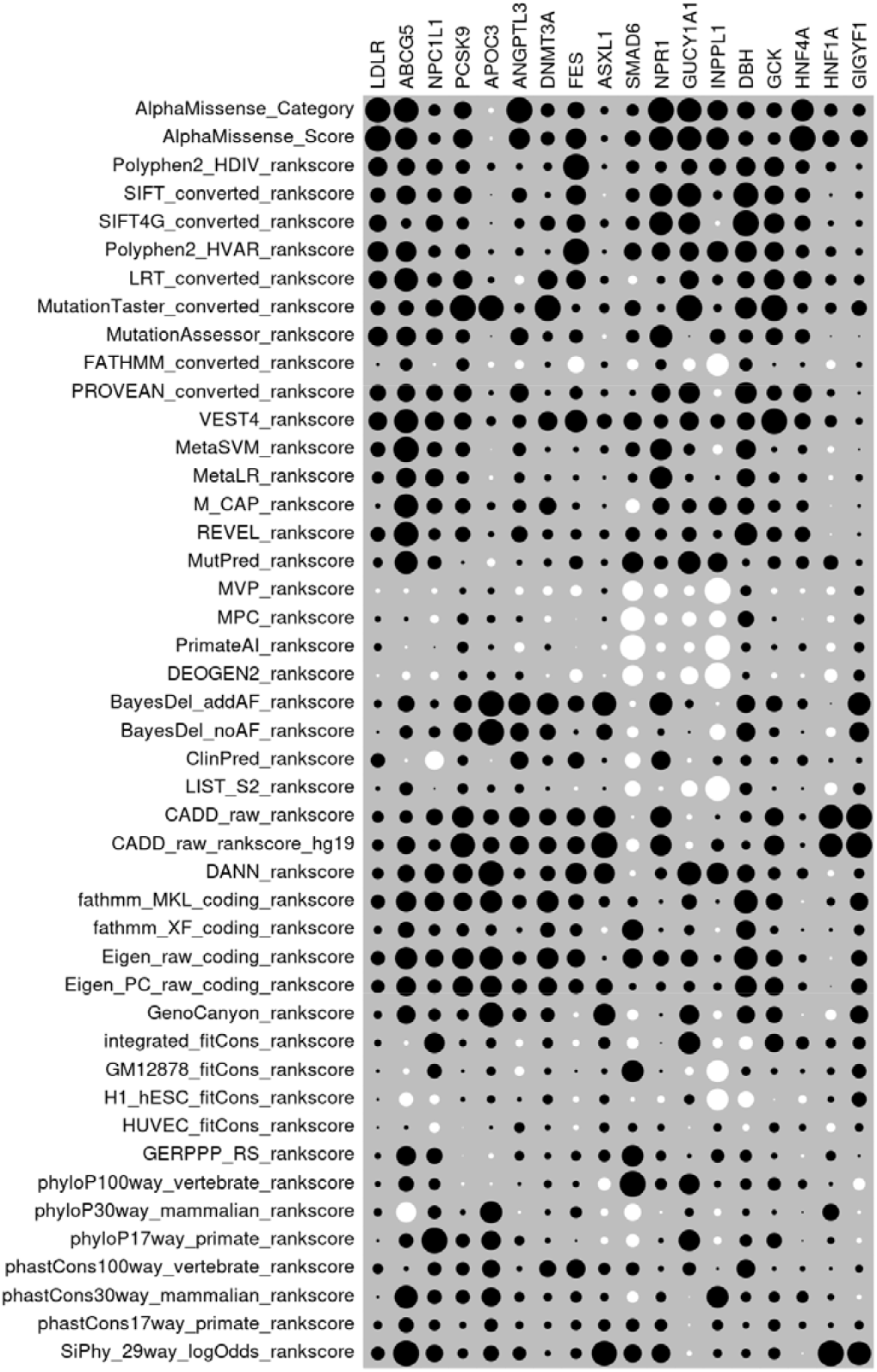
Heatmap of SLPs produced by each secondary annotation for each gene. The sizes of the dots for each gene are proportional to the SLP for each annotation relative to the maximum SLP produced by any annotation for that gene. White circles indicate negative SLPs.

**Table 4.** SLPs produced individually by each secondary annotation from AlphaMissense and dbNSFP for each gene, including sex and principal components as covariates. SLPs of 3 or more are shown in bold and SLPs of 6 or more in bold italics. The final column shows the mean SLP achieved by each annotation across all genes.

| Annotation | *LDLR* | *ABCG5* | *NPC1L1* | *PCSK9* | *APOC3* | *ANGPTL3* | *DNMT3A* | *FES* | *ASXL1* | *SMAD6* | *NPR1* | *GUCY1A1* | *INPPL1* | *DBH* | *GCK* | *HNFA4* | *HNFA1* | *GIGYF1* | Mean |
| --- | --- | --- | --- | --- | --- | --- | --- | --- | --- | --- | --- | --- | --- | --- | --- | --- | --- | --- | --- |
| AlphaMissense Category | **76.51** | 2.07 | 0.72 | **9.76** | -0.45 | **3.89** | 2.90 | 2.97 | 0.41 | 0.31 | **4.82** | **4.38** | 1.20 | 2.57 | **6.49** | **6.15** | 2.28 | 1.12 | **7.12** |
| AlphaMissense Score | **80.33** | 1.62 | 0.74 | **11.70** | -0.15 | 2.56 | 2.93 | **4.32** | 0.30 | 0.53 | **4.15** | **4.35** | 1.46 | 1.83 | **4.36** | **8.24** | **3.73** | 2.06 | **7.50** |
| Polyphen2 HDIV rankscore | **43.87** | 1.04 | 1.34 | **6.14** | 1.04 | 0.25 | 1.08 | **6.87** | 0.18 | 0.36 | 0.96 | 1.89 | 0.82 | **3.04** | **10.61** | 2.36 | 1.33 | 0.26 | **4.64** |
| SIFT converted rankscore | **27.02** | 1.22 | 1.05 | **9.94** | 0.04 | 1.29 | 0.56 | **4.04** | -0.05 | 0.40 | **3.67** | **4.18** | 0.23 | **5.12** | **10.07** | 2.74 | 0.15 | 0.39 | **4.00** |
| SIFT4G converted rankscore | **36.14** | 0.34 | 1.49 | **4.72** | 0.08 | 0.57 | **3.51** | **3.50** | 0.48 | 0.41 | **3.97** | **3.44** | -0.06 | **5.56** | **11.13** | **3.08** | 0.45 | 0.32 | **4.40** |
| Polyphen2 HVAR rankscore | **49.94** | 1.41 | 0.78 | **7.00** | 0.51 | 0.39 | 0.44 | **6.82** | 0.13 | 0.64 | 2.42 | 2.70 | 1.29 | **3.88** | **10.20** | 2.19 | 1.46 | 0.37 | **5.14** |
| LRT converted rankscore | **39.31** | 1.86 | 1.10 | **11.33** | 0.71 | -0.48 | **5.83** | **3.83** | 0.46 | -0.16 | 0.49 | 2.65 | 0.32 | 2.45 | **11.48** | **4.01** | 1.55 | 0.75 | **4.86** |
| MutationTaster converted rankscore | **25.14** | 0.79 | 1.59 | **21.07** | **11.07** | 0.58 | **10.27** | 0.72 | 0.68 | 0.54 | 0.46 | **4.99** | 0.19 | **3.92** | **18.46** | 1.31 | 1.67 | 1.65 | **5.84** |
| MutationAssessor rankscore | **45.73** | 1.28 | 1.43 | **6.21** | 0.00 | 1.85 | 1.53 | 1.94 | -0.12 | 0.37 | **3.69** | 0.00 | 0.69 | 1.65 | **8.19** | 2.67 | 0.00 | 0.00 | **4.28** |
| FATHMM converted rankscore | 0.96 | 0.53 | -0.04 | **5.12** | -0.57 | -0.45 | 0.51 | -2.81 | 0.29 | -0.26 | 0.86 | -1.13 | -1.42 | 1.41 | 0.32 | 0.27 | -1.06 | 0.22 | 0.15 |
| PROVEAN converted rankscore | **31.26** | 1.07 | 1.38 | **9.70** | 0.44 | 1.98 | 0.52 | 1.70 | 0.36 | 0.08 | 2.52 | **3.29** | -0.16 | **3.87** | **6.23** | **4.11** | 0.76 | 0.06 | **3.84** |
| VEST4 rankscore | **40.88** | 1.94 | 1.86 | **9.36** | 1.55 | 1.07 | **5.65** | **5.13** | 1.50 | 0.69 | 1.44 | 2.66 | 0.61 | 2.61 | **18.35** | **3.21** | 1.95 | 0.35 | **5.60** |
| MetaSVM<br>rankscore | **26.19** | 2.14 | 1.16 | **6.59** | -0.01 | 0.99 | 0.46 | 0.31 | 0.47 | 0.44 | **3.40** | 0.95 | -0.27 | **3.23** | 0.91 | 1.03 | -0.55 | 0.00 | 2.64 |
| MetaLR rankscore | **16.92** | 1.32 | 1.69 | **4.89** | -0.27 | 0.64 | 0.70 | 0.36 | 0.08 | 0.18 | **3.63** | 0.49 | 0.06 | 2.75 | **3.16** | 0.96 | -0.28 | 0.35 | 2.09 |
| M CAP rankscore | 2.80 | 1.93 | 1.25 | **7.28** | 1.14 | 1.02 | **4.55** | 0.57 | 0.11 | -0.41 | 2.02 | 1.56 | 0.93 | 2.29 | **4.22** | 1.71 | -0.02 | 0.07 | 1.83 |
| REVEL rankscore | **25.63** | 2.02 | 0.75 | **7.21** | 0.61 | 1.51 | 1.68 | 1.19 | 0.62 | 0.44 | 1.02 | 1.59 | 0.22 | **4.25** | **6.22** | 2.85 | -0.03 | 0.09 | **3.22** |
| MutPred<br>rankscore | **11.04** | 1.77 | 0.98 | 0.36 | -1.08 | 0.19 | 0.96 | 1.92 | 0.32 | 0.93 | 1.37 | **3.80** | 1.11 | 0.48 | **4.65** | 2.04 | 2.94 | 0.31 | 1.89 |
| MVP rankscore | -2.16 | -0.08 | -0.16 | 1.33 | 0.76 | -0.37 | -1.33 | -1.29 | 0.22 | -0.87 | -1.32 | -0.81 | -1.95 | 0.97 | -1.01 | -0.30 | -0.90 | 0.66 | -0.48 |
| MPC rankscore | **3.97** | 0.06 | -0.36 | **3.94** | 1.14 | 0.24 | -0.65 | -0.01 | 0.14 | -1.20 | -1.12 | -1.53 | -0.81 | 2.14 | 0.58 | -0.03 | -0.67 | 0.76 | 0.37 |
| PrimateAI<br>rankscore | **6.96** | -0.02 | 0.01 | **3.93** | 0.55 | -0.56 | 0.60 | -0.01 | -0.20 | -1.34 | -0.63 | -0.80 | -1.77 | 0.74 | 0.93 | 0.70 | -1.06 | 0.75 | 0.49 |
| DEOGEN2<br>rankscore | -0.87 | -0.22 | -0.33 | 2.50 | 1.23 | 0.38 | -0.18 | -1.68 | -0.06 | -0.89 | -0.47 | -2.36 | -1.91 | 0.44 | -0.94 | -0.20 | -2.24 | 0.91 | -0.38 |
| BayesDel addAF<br>rankscore | **7.04** | 0.93 | 0.37 | **9.88** | **11.49** | 2.82 | **7.16** | 2.73 | **3.83** | -0.08 | **3.76** | 0.52 | -0.03 | 2.81 | **7.22** | 1.25 | 0.02 | **3.70** | **3.63** |
| BayesDel noAF<br>rankscore | 0.72 | 0.57 | 0.66 | **11.21** | **11.70** | 1.95 | **3.86** | 0.23 | 1.70 | -0.26 | 0.34 | 0.04 | -0.72 | 2.89 | 2.68 | 0.09 | -1.23 | 2.77 | 2.18 |
| ClinPred<br>rankscore | **24.08** | -0.03 | -1.80 | **3.42** | -0.06 | 1.86 | 1.64 | 2.75 | 0.17 | -0.51 | 2.61 | -0.14 | 0.25 | 0.97 | 1.66 | 1.41 | 0.28 | 0.25 | 2.16 |
| LIST S2 rankscore | 2.52 | 0.60 | 0.00 | **3.53** | 1.06 | 0.48 | 0.28 | -0.15 | 0.08 | -0.52 | -0.15 | -1.99 | -1.75 | 1.28 | 0.53 | 0.08 | -2.11 | 1.05 | 0.27 |
| CADD raw<br>rankscore | **15.96** | 0.57 | 1.42 | **14.65** | **4.28** | 2.40 | **4.40** | **3.46** | **3.22** | -0.02 | **3.28** | -0.22 | 0.07 | 1.12 | **9.53** | 0.53 | **7.81** | **4.72** | **4.29** |
| CADD raw<br>rankscore hg19 | **15.91** | 1.08 | 0.75 | **18.71** | **4.65** | 2.07 | **4.56** | **3.12** | **4.50** | -0.35 | **3.27** | -0.39 | 0.47 | 0.65 | **11.03** | 0.27 | **7.36** | **4.79** | **4.58** |
| DANN rankscore | **15.69** | 1.00 | 1.98 | **12.04** | **11.03** | 0.74 | **3.58** | **4.59** | 2.89 | -0.07 | 1.02 | **4.20** | 1.36 | 2.55 | **4.32** | 1.51 | -0.51 | 0.75 | **3.82** |
| fathmm MKL<br>coding rankscore | **23.47** | 1.39 | 1.95 | **10.90** | **8.47** | 1.14 | **7.19** | 2.44 | 0.79 | 0.21 | 0.13 | 1.72 | 0.05 | **4.59** | **7.75** | -0.09 | 0.88 | 2.33 | **4.18** |
| fathmm XF coding<br>rankscore | **7.74** | 0.89 | 0.67 | **4.86** | 2.09 | 0.19 | **3.42** | 1.52 | -0.24 | 0.94 | 0.15 | 0.75 | -0.06 | **3.10** | 1.76 | 0.09 | 0.18 | 0.77 | 1.60 |
| Eigen raw coding<br>rankscore | **23.53** | 1.65 | 1.66 | **14.31** | **9.30** | 1.07 | **6.98** | **3.34** | 0.12 | 0.82 | 1.78 | 2.13 | 0.15 | **4.52** | **7.71** | 0.74 | -0.07 | 1.78 | **4.53** |
| Eigen PC raw<br>coding rankscore | **21.12** | 1.28 | 1.03 | **12.89** | **7.87** | 1.28 | **5.20** | 2.29 | 1.82 | 0.11 | 0.66 | 0.93 | 0.19 | **3.92** | **9.27** | 0.89 | 0.01 | 1.78 | **4.03** |
| GenoCanyon<br>rankscore | **7.77** | 1.16 | 0.79 | **4.33** | **10.23** | 1.08 | 2.98 | -0.24 | **3.24** | -0.25 | 0.07 | 2.89 | -0.21 | 2.67 | **7.38** | -0.03 | -1.54 | 2.35 | 2.48 |
| integrated fitCons<br>rankscore | **5.69** | -0.07 | 2.13 | 1.63 | 1.74 | -0.31 | 1.63 | -0.45 | 0.95 | -0.28 | 0.00 | **3.62** | -0.28 | -1.52 | **9.51** | 2.04 | 1.35 | 1.17 | 1.59 |
| GM12878 fitCons<br>rankscore | 1.21 | -0.12 | 1.07 | 0.29 | 1.39 | -0.51 | 0.72 | 0.07 | 0.23 | 1.02 | 0.06 | -0.52 | -1.40 | 0.68 | 1.19 | 0.25 | 0.74 | 1.44 | 0.43 |
| H1 hESC fitCons<br>rankscore | 0.65 | -0.65 | -0.47 | 0.65 | 1.11 | 0.11 | 1.51 | -0.35 | 0.63 | -0.04 | -0.37 | 0.76 | -1.36 | -2.12 | -0.05 | -0.81 | 0.53 | 1.56 | 0.07 |
| HUVEC fitCons<br>rankscore | 0.57 | 0.06 | -0.53 | -0.01 | 0.24 | 0.63 | 0.49 | -0.28 | 0.66 | 0.17 | 0.36 | -0.03 | 0.19 | -0.37 | 2.70 | 0.60 | -1.03 | 0.50 | 0.27 |
| GERPPP RS<br>rankscore | **4.59** | 1.30 | 1.33 | -0.01 | -0.46 | 0.52 | 0.67 | 0.99 | 0.94 | 0.98 | 0.76 | 0.15 | 0.50 | 1.16 | 1.04 | -0.11 | 1.31 | 0.05 | 0.87 |
| phyloP100way<br>vertebrate<br>rankscore | **3.85** | 0.79 | 0.68 | -0.26 | -0.12 | 0.07 | 0.90 | 0.42 | -1.07 | 1.42 | 1.00 | **3.15** | 0.14 | 1.05 | **4.61** | 0.98 | 0.23 | -1.01 | 0.94 |
| phyloP30way<br>mammalian<br>rankscore | **7.68** | -1.35 | 0.90 | 0.87 | **8.54** | -0.04 | 0.49 | 1.37 | -0.35 | -0.60 | -0.04 | 0.56 | -0.14 | 0.42 | 0.64 | 0.02 | **3.77** | -0.07 | 1.26 |
| phyloP17way<br>primate<br>rankscore | 0.80 | 0.69 | **3.44** | **6.09** | **6.55** | 0.56 | 0.47 | 0.05 | -0.35 | -0.36 | 0.18 | **3.38** | -0.17 | 1.24 | **6.41** | 0.02 | 0.65 | -0.17 | 1.64 |
| phastCons100way<br>vertebrate<br>rankscore | **12.90** | 0.08 | 0.96 | **5.64** | **5.63** | 0.18 | **4.37** | **3.67** | 0.97 | 0.37 | 0.23 | 1.31 | 0.02 | 2.87 | 0.57 | 0.35 | 0.17 | 0.40 | 2.26 |
| phastCons30way<br>mammalian<br>rankscore | 0.79 | 1.80 | 1.06 | **6.19** | **6.39** | 0.58 | 1.06 | 1.30 | 0.46 | -0.27 | 0.76 | -0.01 | 1.40 | 0.87 | **3.33** | 1.71 | 0.58 | -0.15 | 1.55 |
| phastCons17way<br>primate<br>rankscore | **8.83** | 0.77 | 0.63 | 1.45 | **3.04** | 0.36 | 0.81 | 1.09 | 0.99 | 0.37 | 1.10 | -0.03 | 0.34 | 0.34 | **3.99** | 0.65 | 0.42 | 0.57 | 1.43 |
| SiPhy 29way<br>logOdds<br>rankscore | **21.86** | 2.25 | 1.54 | **3.43** | **4.30** | 1.05 | 0.51 | 1.25 | **4.24** | 0.69 | 2.50 | -0.18 | 0.22 | 0.69 | **3.90** | -0.02 | **9.05** | **4.05** | **3.41** |

When a secondary annotation method is able to produce a high SLP, there is inconsistency between genes with regard to the relative performance of the different methods. While the AlphaMissense annotations have the best performance on average across all genes (for AlphaMissense category SLP = 7.12 and for AlphaMissense score SLP = 7.50), they actually produce the maximum SLP for only 4 genes: *LDLR, ANGPTL3, NPR1* and *HNFA1*. There are some genes where AlphaMissense is able to produce reasonable evidence for association but other methods do considerably better. For example, *PCSK9* yields SLP = 11.70 with the AlphaMIssense score but SLP = 21.07 with MutationTaster, while *GCK* yields SLP = 6.49 with the AlphaMissense category but SLP = 18.46 with MutationTaster and SLP = 18.35 with the Variant Effect Scoring Tool (VEST4) (Carter et al., 2013). More strikingly, there were genes for which AlphaMissense was not able to find any evidence for association whereas another method performed well. For example, AlphaMissense produces negligible SLPs for both *APOC3* and *ASXL1* whereas MutationTaster produces SLP = 11.07 for *APOC3* and CADD produces SLP = 4.50 for *ASXL1*.

## Discussion

Examining this relatively small number of gene-phenotype pairs in detail is sufficient to establish that there is dramatic variability in the performance of secondary annotation methods in terms of their ability to produce evidence for association. This would seem to have a number of implications.

The first implication is that it is not at all clear what is the optimal approach to use when testing for association between coding variants and a complex phenotype. There are choices to be made between carrying out multiple different analyses using different categorisations, annotation methods and weighting schemes or attempting to combine information from multiple sources into a smaller number of analyses. The results shown here seem to demonstrate that relying on a single annotation method would risk failing to detect some real associations, although if one were forced to rely on a single method then it does seem that AlphaMissense has the best performance on average.

The second implication seems to be that, because different methods work better for different gene-phenotype pairs, one would want to take account of this if the aim was to use sequence data for individual level risk prediction. For example, if one wished to obtain a comprehensive assessment of an individual’s risk of developing type 2 diabetes then based on these results one might use MutationTaster to classify *GCK* variants, AlphaMissense for *HNF4A* and CADD for *HNF1A* and for *GIGYF1*. It would be suboptimal to apply a single annotation method to characterise variants across multiple genes.

Finally, it seems that it would be very desirable to be in a position where one could in advance identify for a given gene or gene-phenotype pair which annotation method would best distinguish the relevant variants. As knowledge accrues it would be helpful to investigate what are the characteristics of a gene which mean that one method will perform well and another poorly. Ultimately one would then seek to develop an automated method in which the first step was gene classification and then this would be followed by application of a gene-relevant annotation method.

## Acknowledgments

This research has been conducted using the UK Biobank Resource. The author wishes to acknowledge the staff supporting the High Performance Computing Cluster, Computer Science Department, University College London. The author wishes to thank the participants who volunteered for the UK Biobank project.

## Ethics statement

UK Biobank had obtained ethics approval from the North West Multi-centre Research Ethics Committee which covers the UK (approval number: 11/NW/0382) and had obtained written informed consent from all participants. The UK Biobank approved an application for use of the data (ID 51119) and ethics approval for the analyses was obtained from the UCL Research Ethics Committee (11527/001).

## Data availability

The raw data is available on application to UK Biobank. Detailed results with variant counts cannot be made available because they might be used for subject identification. Scripts and relevant derived variables will be deposited in UK Biobank. Software and scripts used to carry out the analyses are also available at https://github.com/davenomiddlenamecurtis.

## Author contributions

DC carried out the analyses and prepared the manuscript.

